# Mechanistic Assessment of Norepinephrine Therapy versus Angiotensin-II on Organ Function in Septic Shock (MANTRA): Study Protocol for a multicenter open-label randomized trial

**DOI:** 10.64898/2026.09.12.26362906

**Authors:** Michael R. Filbin, Daniel E. Leisman, Ana Pachano-Bravo, Marcia B. Goldberg, Kathryn Hibbert, Olivia Nelson, Benjamin Rappaport, Simon A. Mahler, Lynnette Harris, Estelle B. Besong, Brandon Reeves, Ryan C. Maves, Andrew Petrilli, Christopher L. Schaich, D. Clark Files, Kevin Gibbs, Mark C. Chappell, Ashish K. Khanna

## Abstract

**Introduction:** Norepinephrine remains first-line vasopressor therapy for septic shock despite its association at high doses with undesirable cardiovascular, kidney, and immune effects. Angiotensin-II (Ang-II) is a non-catecholamine vasopressor of the renin-angiotensin-aldosterone system (RAAS) now approved to increase blood pressure in vasodilatory shock. RAAS dysregulation is common in septic shock and may represent a treatable trait. However, few prospective investigations evaluate the effect of early Ang-II use in septic shock on dysfunctional RAAS biology, host-response, or clinical outcomes. Therefore, we designed a mechanistic clinical trial to test the hypothesis that early Ang-II therapy in septic shock normalizes systemic RAAS abnormalities and modulates innate immune responses.

**Methods and Analysis:** The **M**echanistic **A**ssessment of **N**orepinephrine **T**he**R**apy vs. **A**ngiotensin-II in Septic Shock (MANTRA) trial is an investigator-initiated, prospective, multicenter, open-label, randomized mechanistic clinical trial. Adults with septic shock by Sepsis-3 criteria receiving a norepinephrine-equivalent dose (NED) of 0.10-0.50 mcg/kg/min will be randomized 1:1 to primary Ang-II (to a maximum 40 ng/kg/min) or norepinephrine (to a maximum 35 mcg/min) treatment strategies over a 48-hour period. The primary outcome is the between-group difference in plasma renin trajectory from baseline to 24 hours represented as a treatment-by-time interaction term in a linear mixed-effects model. With n=78, the trial has 90% power (at two-sided α=0.05) to detect a 25% between-group difference in 24-hour renin. Secondary clinical outcomes include longitudinal NED, hours alive and vasopressor-free at 72 hours, a 28-day composite outcome (days alive and free of vasopressors, renal replacement therapy, and mechanical ventilation), treatment failure requiring crossover or protocol discontinuation, and thrombotic and arrhythmia events through 28 days. Secondary mechanistic outcomes include serial RAAS biomarkers (prorenin, soluble prorenin receptor, angiotensinogen, ACE, ACE2, Ang-II, angiotensin-(1–7), aldosterone, and DPP3), immune measures (ex vivo monocyte function, monocyte gene expression, plasma cytokines), and organ injury biomarkers. Enrollment is planned from 11/2026-7/2030.

**Ethics and Dissemination:** The study has been approved by the Advocate Health/Wake Forest University School of Medicine ethics commitee (IRB00115287). Informed consent is required. Results will be submitted for publication in peer-reviewed journals and presented at national and international scientific meetings, regardless of the direction of findings. Results will be posted to ClinicalTrials.gov.

**Trial Registration:** Clinicaltrials.gov - NCT06746753

**Administrative Information:** This manuscript has been written in accordance with the SPIRIT (Standard Protocol Items: Recommendations for Interventional Trials) guidelines. The administrative information is described in **Table 1**.

**Protocol Version:** Protocol Version 2, 11-08-2026 – NCT06746753

**OPEN SCIENCE:** *Trial Registration:* Mechanistic Assessment of Norepinephrine Therapy vs. Angiotensin-II in Septic Shock (MANTRA); Trial identification number NCT06746753; date of registration: 03-31-2026. https://clinicaltrials.gov/study/NCT06746753

*Data Sharing:* Data will be available from the corresponding author on reasonable request.

*Funding and conflicts of interest:* Sources of funding and support: National Institutes of Health (NIH) National Heart Lung Blood Institute (NHLBI) R01HL177834 (U.S. NIH Grant/Contract) Dissemination Policy

*Dissemination Policy:* Key findings will be published in peer-reviewed journals, and presented at national and international scientific meetings, ensuring confidentiality of individual participants.

*Strengths and limitations of this study:*

- The active-comparator design tests angiotensin-II against norepinephrine under identical protocolised MAP targets, allowing the comparison of drug-specific RAAS modulatory effects independent of differential effects on blood pressure restoration.
- Serial sampling profiles RAAS peptides, innate immune function, and organ-injury biomarkers facilitates mechanistic evaluation of how vasopressor strategy shapes the host response within a randomized clinical experiment.
- Although the sample size is adequate for the biomarker primary endpoints and to establish feasibility for a larger trial, it does not provide sufficient power for hard clinical endpoints, which remain exploratory.
- Clinicians retain discretion over background and rescue vasopressors, steroids, and treatment crossover, which may attenuate differences between-arms.
- The 48-hour intervention window limits inference about clinician-assessed outcomes and the durability of treatment effect.

## INTRODUCTION

### Background and Rationale

Sepsis is life-threatening organ dysfunction caused by a dysregulated host-response to infection.^1^ Septic shock is characterized by severe hypotension, typically due to inappropriate vasodilation, and hypoperfusion of key organs. Other than early antibiotic administration, sepsis interventions, including drugs that modulate inflammation, have not proven beneficial. New therapeutic approaches are clearly needed, and because sepsis is heterogeneous, rapid identification of patients at risk of worse outcomes or likely to respond to specific therapeutic interventions would be beneficial. Recent studies have shown that activation of the renin-angiotensin-aldosterone system (RAAS) occurs early in septic shock, and elevated circulating renin, the early and committed step in the overall RAAS cascade, is a better predictor of adverse outcomes than lactate.^2–6^ However, downstream RAAS signaling and angiotensin-II generation are commonly dysfunctional in sepsis, which contributes not only to hypotension but possibly also to immune dysfunction^7–9^ that may be exacerbated by putative immunosuppressive effects of norepinephrine (NE),^10–12^ the most used vasopressor. Among patients with refractory vasodilatory shock and elevated baseline renin in the Angiotensin-II in High Output Shock (ATHOS-3) trial, angiotensin-II (Ang-II) treatment was associated with improved survival versus placebo.^2,13–15^ In murine sepsis models, Ang-II improved immune function and bacterial clearance.^16^ These observations suggest that RAAS dysregulation could represent a treatable trait in septic shock and that Ang-II, in addition to increasing blood pressure, could benefit patients with septic shock through direct immunologic effects. We therefore designed the **M**echanistic **A**ssessment of **N**orepinephrine **T**he**R**apy vs. **A**ngiotensin-II in Septic Shock (MANTRA) trial to test the hypotheses that, in early septic shock, primary vasopressor therapy with Ang-II would ameliorate RAAS dysfunction as primarily measured by plasma renin trajectories, and would modulate the host response versus standard care with norepinephrine. The trial was peer reviewed and approved as part of the funded NIH/NHLBI grant R01HL177834.

### Objectives

We hypothesize that a primary Ang-II vasopressor approach will more effectively normalize plasma renin compared to the standard primary NE approach and that the serial comparison of these two approaches will additionally generate relevant mechanistic insights into the biology of septic shock.

**- Primary Objective:**

- Evaluate the effect of Ang-II predominant versus NE predominant treatment on plasma renin levels after 24 hours of therapy in early septic shock.
**- Secondary Objectives:**

- Characterize the effect of Ang-II versus NE treatment on the trajectory of markers of RAAS function and organ injury.
- Identify which circulating RAAS biomarkers are most strongly associated with favorable treatment response to Ang-II therapy.
- Interrogate characteristics and mechanisms of innate immune modulation with Ang-II versus NE.

## METHODS: Trial Design

### Trial Design

The MANTRA study is an investigator-initiated, parallel, prospective, three-center, open label, randomized, mechanistic clinical trial. Adult patients with septic shock will be randomly assigned to a treatment strategy using primarily Ang-II vs. NE for up to 48 hours. The expected duration of the subject’s participation is 4 weeks after inclusion; the study ends after the day-28 assessment.

### CONSORT Diagram

The Consolidated Standards of Reporting Trials (CONSORT) diagram is shown in **Figure 1**.

**Figure 1.**
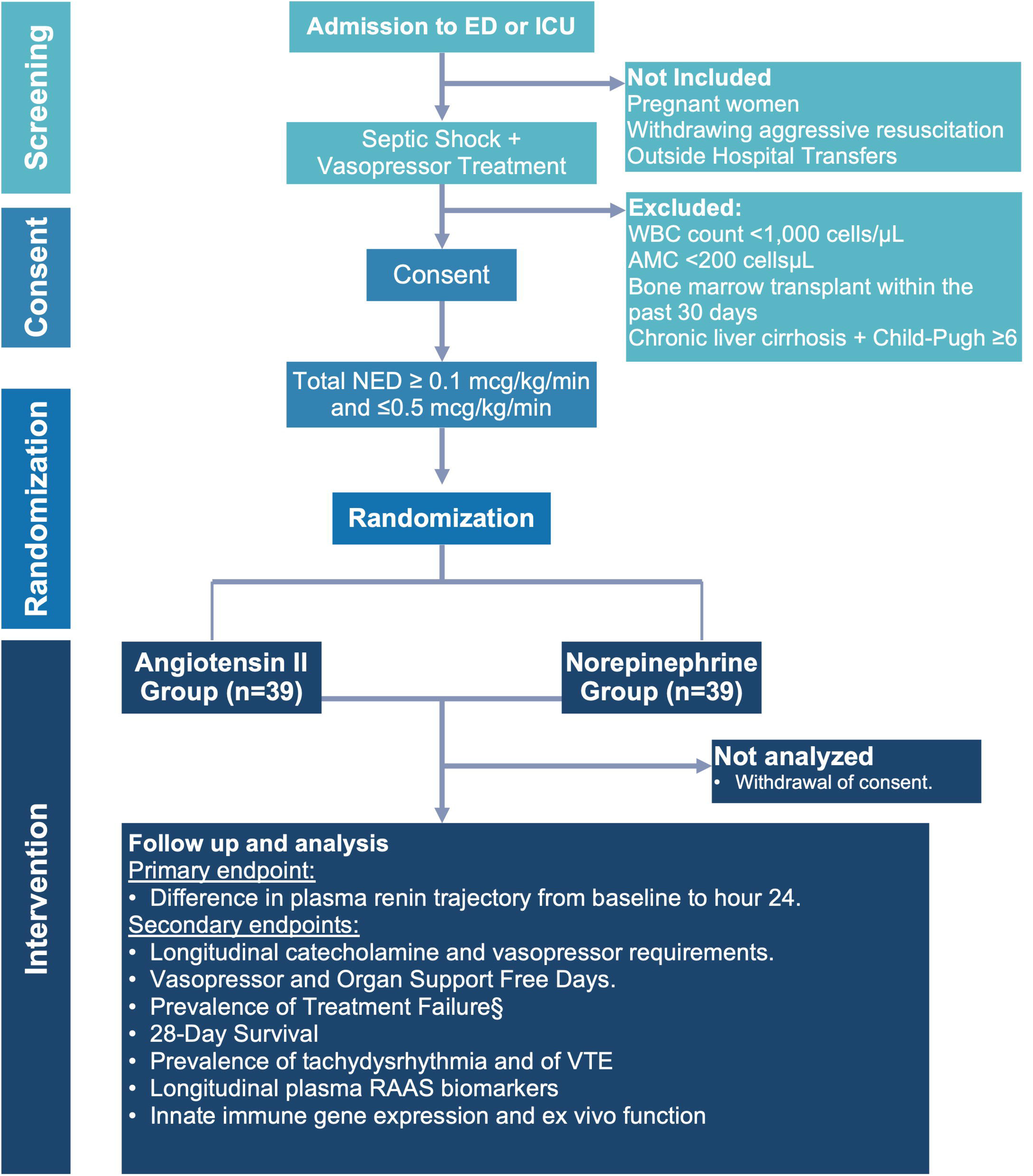
Consort Diagram for MANTRA Trial. Abbreviations: ED: Emergency Department, ICU: Intensive Care Unit, NE: Norepinephrine, NED: Norepinephrine Equivalent Dose, VTE: Venous Thromboembolism, RAAS: Renin-Angiotensin-Aldosterone System. ^§^Treatment Failure: Defined as inability to meet target MAP despite maximum protocolized dose of assigned primary vasopressor (Ang-II or NE).

## METHODS: Participants, Interventions and Outcomes

### Trial Setting

The MANTRA trial will take place in the emergency departments (EDs) and intensive care units (ICUs) at Atrium Health Wake Forest Baptist Medical Center (Winston-Salem, NC), Atrium Health High Point Medical Center (High Point, NC), and the Massachusetts General Hospital (Boston, MA).

### Eligibility Criteria

#### Inclusion Criteria

We will include adult patients aged 18 years and older presenting to the ED or ICU who meet the following criteria:

**(1)** Septic shock, defined by Sepsis-3 criteria: *(1a)* suspected or known infection, *(1b)* hypotension requiring vasopressors, and *(1c)* lactate >2 mmol/L;
**(2)** Receipt of either norepinephrine or phenylephrine with or without additional vasopressor support;
**(3)** A total norepinephrine equivalent dose (NED) ≥0.1 mcg/kg/min and ≤0.5 mcg/kg/min;
**(4)** Provision of informed consent by the patient or a legally authorized representative (LAR) and the ability to randomize within 24 hours of first reaching the NED threshold and within 48 hours of hospital arrival.

Informed consent from the patient or their LAR may be obtained after initial fluid resuscitation and vasopressor initiation per standard practice and meeting inclusion criteria 1, 2, and 4 above, after which the patient will be further screened for study eligibility. To be eligible for randomization, patients must be within the vasopressor NED dosing range listed in inclusion criterion 3 above and not meet any exclusions listed below.

#### Exclusion Criteria

We will exclude patients with any of the following criteria: younger than 18 years old; urgent surgery is anticipated; leukocyte count <1,000 cells/μL; absolute monocyte count <200 cells/μL; history of hematopoietic stem cell transplant within the past 30 days; history of liver cirrhosis with Child-Pugh score ≥6; pregnant women; patients that will be withdrawing aggressive resuscitation including withdrawal of vasopressor support; external hospital ICU transfers; incarcerated persons; and patients who present any additional concerns to the study investigator that could confound the results or pose an extra risk for them if they participate in the study.

### Intervention and Comparator

#### Interventions

Eligible patients who are within 24 hours of first reaching the NED threshold and within 48 hours of hospital arrival will be randomly assigned in a 1:1 ratio to either the Ang-II group or the NE group. Study procedures will be conducted in two phases (**Figure 2**) during the 48-hour intervention period (Initiation & Steady state) with de-escalation occurring during the 48h intervention if shock improves.

1. **Initiation**: Study drug will be initiated and up-titrated to an increased MAP target of 75-80 mmHg while other vasopressors already infusing (background vasopressors) are down-titrated if applicable. Both arms will implement the same increased MAP goal for this period to ensure the consistency of hemodynamic targets between treatment groups. The initiation phase will be for 1 hour starting at hour 0 with the goal to achieve a steady state of study drug administration maintaining MAP ≥75 mmHg without vasopressor titration.

a. **Ang-II arm**: Initiate Ang-II at 5 ng/kg/min. Up-titrate Ang-II while down-titrating background vasopressor to maintain MAP ≥75 mmHg. The goal is to maximize Ang-II dose and minimize (optimally discontinue) background vasopressors.
b. **NE arm**: Continue NE, or initiate NE if the patient is primarily receiving phenylephrine, and titrate to achieve MAP ≥75 mmHg. For subjects receiving phenylephrine at baseline, down-titrate phenylephrine while up-titrating NE.
2. **Steady state**: After the 1-hour initiation phase, the MAP goal reverts to ≥65 mmHg (or other hemodynamic target if specified by the clinical team) and study drug infusion is managed by the clinical team for the duration of the study period. If the target MAP cannot be achieved with the assigned vasopressor’s maximum study dose (listed below), the patient will be designated a treatment failure. In both arms, the use of adjunct vasopressors other than Ang-II or NE (e.g., vasopressin, epinephrine, etc.), or the substitution of phenylephrine for norepinephrine, and/or the addition of stress-dose steroids will be at the discretion of the clinical team.**De-escalation if shock improves**: If the patient clinically improves during the 48h intervention period, then vasopressors can be weaned by the clincal team.

a. **Ang-II arm**: Background vasopressors shall be weaned first, and study Ang-II last. If at the end of the 48-hour intervention period the patient still requires vasopressor support, study Ang-II will be weaned off, and other vasopressors titrated as needed to maintain a MAP of 65 mmHg or another hemodynamic target set by the clinical team. After the 48h hour intervention period, the clinical team may initiate clinical supply of Ang-II if deemed clinically indicated.
b. **NE arm**: Weaning strategy shall be at the discretion of the clinical team. For patients receiving both vasopressin and NE, the recommendation will be to wean the vasopressin first, followed by the NE. On the other hand, for patients that still require vasopressor support at the end of the 48-hour study period, the NE infusion may be continued per usual clinical management procedures.

**Figure 2.**
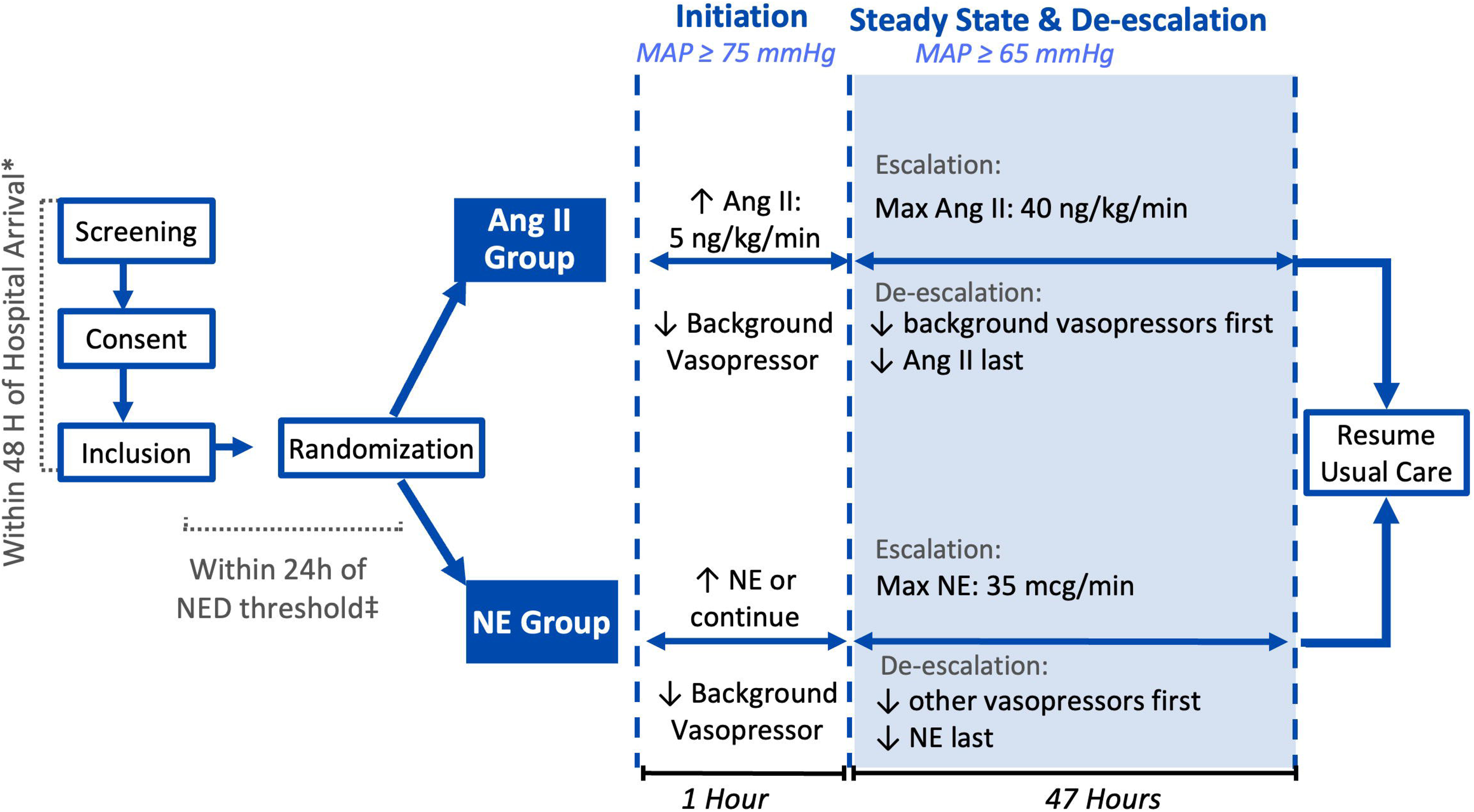
Study Design: Angiotensin II group vs. Norepinephrine Group. Schematic representation of the study design, including participant inclusion, allocation, and group-specific interventions throughout the trial. *Participants will be eligible for randomization within 48 hours of hospital arrival‡ Randomization will occur within 24 hours after participants first reaches the NED threshold. Abbreviations: Ang-II: Angiotensin II; NE: Norepinephrine.

Upon completion of the 48-hr intervention period, if the subject is in the Ang-II arm and still receiving study Ang-II, then Ang-II will be weaned to off while background vasopressors are titrated up as dictated by the clinical team. If the subject is in the NE arm and still receiving NE, then clinical NE will be continued at the discretion of the clinical team as per usual care.

#### Criteria for discontinuing or modifying allocated interventions

There are no formal stopping rules given the specified dose ranges of Ang-II vs NE in each arm and the flexibility for the clinical team to maintain or add adjuvant circulatory support in addition to the study vasopressor. At any point during the 48-hour intervention period, the clinical team or investigator may down-titrate or discontinue study vasopressor if there is an attributable safety concern or side effect.

##### Treatment failure

Treatment failure is defined separately for each intervention group as follows:

- **Ang-II arm**: Maximum steady-state dose of study Ang-II shall not exceed 40 ng/kg/min. If MAP goal cannot be maintained with this maximum dose and permitted adjuncts, all rescue vasopressor support will be at the discretion of the clinical team. Ang-II will be maintained at 40 ng/kg/min throughout the remainder of the intervention period unless the study team wishes to titrate it down or discontinue it.
- **NE arm**: Maximum steady state dose of study NE shall not exceed 35 mcg/min. If MAP goal cannot be maintained with this maximum dose and permitted adjuncts, all rescue vasopressor support will be at the discretion of the clinical team. This support may include exceeding the 35 mcg/min maximum dose of NE and/or the addition of clinical supply Ang-II at the full discretion of the clinical team.

As detailed above, in the event of hemodynamic instability despite maximum dose of study vasopressor, adjunctive therapy, dose modification, discontinuation, or alteration of the treatment strategy is at the discretion of the clinical team. Treatment failure events will be recorded and included in secondary outcomes analysis (see below).

#### Strategies to improve adherence to intervention protocols

ED and ICU staff will be trained in advance on study procedures and titration goals in the Ang-II vs NE arms. Most importantly, just-in-time training will be performed with clinical staff caring for enrolled patients. There will be frequent communication between the investigator team and clinical staff before and during the 48-hour intervention period to ensure protocol understanding and adherence. Vasopressor infusion initiation, dosing, dose changes, and discontinuation is routinely documented by nursing staff in the electronic medical record (Epic at both participating institutions). Protocol adherence will be ensured through frequent (at least every 8 hours) review of vasopressor administration documentation, discussion with clinical staff, and assessment for adverse events.

Research staff will communicate with participants and/or their families throughout the duration of the study to ensure protocol understanding, participant retention and adherence to study visit follow-ups. If discharged, participants will be contacted by telephone for the 7-day and 28-day post-intervention follow-up visits, and reminder calls will be used as needed to promote completion of outcome assessments and minimize loss to follow-up.

#### Relevant concomitant care and interventions that are permitted or prohibited during the trial

All standard clinical care and usual supportive therapies will be permitted during the study period, including use of glucocorticoids and adjuvant or rescue vasopressors as is detailed above in study procedures. The use of thromboprophylaxis will be at the discretion of the clinical team and per usual care. Modifications to the protocolized titration of Ang-II vs NE will be allowed when deemed clinically necessary and will be documented accordingly.

### Outcomes

#### Primary Outcome

The primary outcome is the between-group difference in plasma renin trajectory from baseline to 24 hours between intervention arms represented as a treatment-by-time interaction term in a linear mixed-effects model, as a measure of restoration of RAAS homeostasis.

We selected renin level as the primary outcome because renin performs at least as well as lactate as a marker of malperfusion^4–6^ and its release is driven by both RAAS dysregulation and hypoperfusion. Therefore, the previously reported fall in renin and differential benefit of Ang-II treatment in the ATHOS-3 trial (a placebo-controlled trial with background vasopressors fixed for the initial 3 hours) could conceivably reflect a bystander effect of non-specific vasoplegia treatment rather than a drug-specific effect of exogenous Ang-II on endocrine pathology.^2^ By evaluating plasma renin trajectory in this study with Ang-II treatment versus a norepinephrine comparator, we address this previous limitation of the literature directly.

#### Secondary Outcome

Secondary outcomes will evaluate the association between circulating RAAS biomarkers and disease severity, as well as the relationship of treatment arm with clinical and biological outcomes. For this we will be assessing:

- Longitudinal vasopressor requirement (norepinephrine equivalent dose [NED]) at 0 to 3, 12, 24, 48 and 72h.
- Hours alive and vasopressor-free within 72 hours of randomization to study drug.
- A composite outcome of days alive, vasopressor-free days, renal replacement therapy (RRT)-free days, and mechanical ventilation (MV)-free days out to 28 days.
- Number of patients meeting the above predefined criteria for treatment failure.
- Routine clinical measurements:

- *Severity of illness*: SOFA, APACHE-II scores.
- *Indicators of organ dysfunction*: Circulatory - lactate, vasopressor requirements (NED); renal - Kidney Disease Improving Global Outcomes AKI stage, creatinine, BUN, bicarbonate; respiratory - presence of ARDS, hypoxemia severity (SpO2/FiO2 [S/F] ratio); hepatic - serum aspartate and alanine aminotransferases, alkaline phosphatase, bilirubin, albumin. Biomarkers of organ injury - troponin-T, neutrophil gelatinase-associated lipocalin, kidney injury molecule-1, soluble receptor for advanced glycation end products, angiopoietin-2, syndecan-1, protein C, S-100β.
- *Serum markers of systemic inflammation*: Inflammatory cytokines -TNFα, sTNFR1, IL-6, sIL-6R, IL-8, IL-1β, IFN-, IL12p70, IL-17, MIP1-α/β, IP-10; anti-inflammatory cytokines - IL-10, IL-1RA, IL-12p40; other inflammation-associated markers include platelet count, and absolute counts of neutrophils, granulocytes, lymphocytes, and the neutrophil-lymphocyte ratio.
- Baseline concentrations, ratios, and longitudinal measurements of plasma levels of RAAS-associated proteins and peptides: prorenin, renin, plasma renin activity (PRA), soluble prorenin receptor (sPRR), aldosterone, intact angiotensinogen, Ang-II, Ang-(1-7), ACE, ACE2, DPP3.
- Innate immune function as measured by ex vivo monocyte function, gene expression and cell signaling events.

#### Safety Outcomes

Intervention-specific safety outcomes will evaluate for the development of thrombotic or cardiac complications, including new deep vein thrombosis (DVT), pulmonary embolism (PE), new-onset atrial fibrillation, new-onset supraventricular tachycardia (excluding sinus tachycardia), hemodynamically significant atrial fibrillation with rapid ventricular response, or other ventricular arrhythmias requiring pharmacologic therapy, need for electrical cardioversion, or cardiopulmonary resuscitation (CPR). Safety outcomes will be evaluated through study day 28. Clinical safety outcomes will be assessed by the study team through a combination of chart review and communication with the primary clinical service. Additionally, the subject or LAR will be contacted on study days 7 & 28 to assess for the occurrence of safety outcomes. Diagnostic testing for DVT/PE and for tachydysrhythmias at the discretion of the clinical team. The clinical use and duration of therapeutic anticoagulation and thromboprophylaxis will be recorded for all patients.

### Harms

In general, the risks involved with this clinical trial are those associated with the vasopressors being compared (Ang-II and NE). Both vasopressors are FDA-approved drugs to increase blood pressure in vasodilatory shock.^17,18^ Their safety profile is detailed within the package insert and their use is not likely to generate an additional risk during this protocol. Nevertheless, there are inherent risks associated with administration of vasopressor agents common to both Ang-II and NE. The trial protocol includes regular monitoring of adverse events, clinical outcomes, and an interim analysis by an independent data and safety monitoring board.

### Participant timeline

The participant timeline is presented in **Table 2**.

**Table 1.** Administrative Information.

|  |  |
|---|---|
| **Title** | **Mechanistic Assessment of Norepinephrine Therapy versus Angiotensin-II in Septic Shock on Organ Function in Septic Shock (MANTRA): Study Protocol for a multicenter open-label randomized trial** |
| **Trial registration** | Trial identification number NCT06746753 03-31-2026. |
| **Protocol version** | IRB00115287 (11-06-2024) |
| **Contact information of study sponsor** | National Institutes of Health (NIH) National Heart Lung Blood Institute (NHLBI)<br>R01HL177834 (U.S. NIH Grant/Contract) |
| **Role of study sponsor** | The NIH provides resources to support the time and necessary materials to conduct this trial. The principal investigator, co-principal investigators, and other researchers are actively involved in the study design, data collection, management, analysis and interpretation. |
| **Role of coordinating center** | Advocate Health-Wake Forest School of Medicine and the Massachusetts General Hospital are involved in the coordination, monitoring and good practices of the clinical trial. |

**Table 2.**
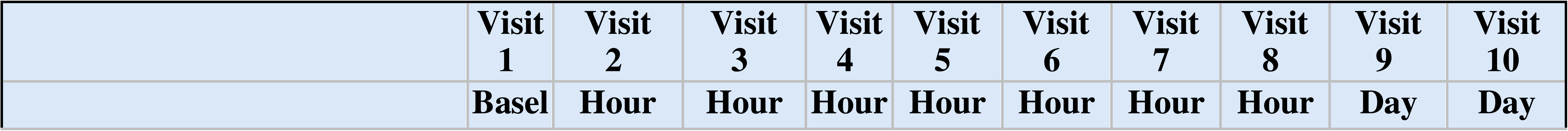
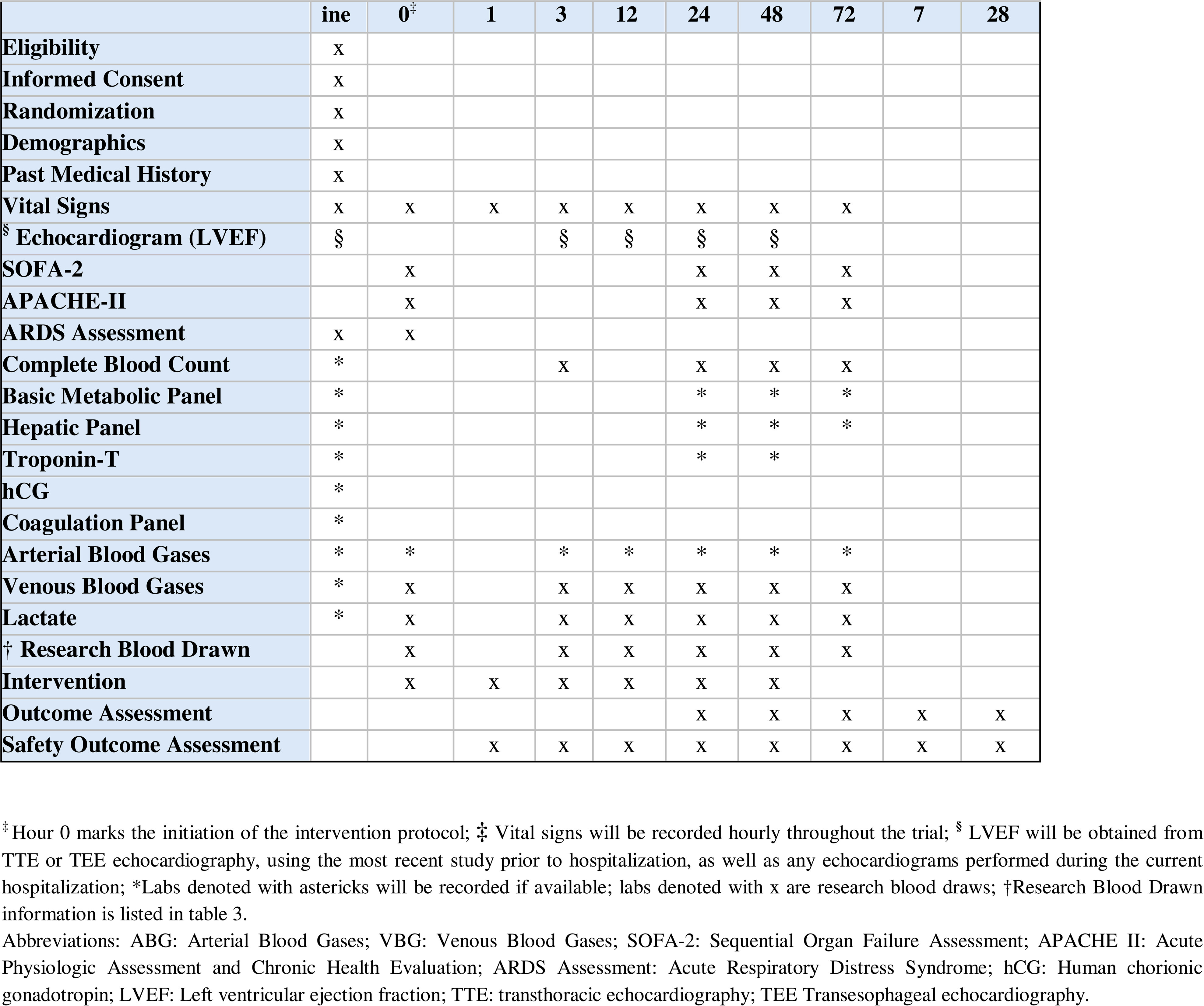
Participant Timeline.

|  | **Visit 1** | **Visit 2** | **Visit 3** | **Visit 4** | **Visit 5** | **Visit 6** | **Visit 7** | **Visit 8** | **Visit 9** | **Visit 10** |
|---|---|---|---|---|---|---|---|---|---|---|
|  | **Baseline** | **Hour 0<sup>‡</sup>** | **Hour 1** | **Hour 3** | **Hour 12** | **Hour 24** | **Hour 48** | **Hour 72** | **Day 7** | **Day 28** |
| **Eligibility** | x |  |  |  |  |  |  |  |  |  |
| **Informed Consent** | x |  |  |  |  |  |  |  |  |  |
| **Randomization** | x |  |  |  |  |  |  |  |  |  |
| **Demographics** | x |  |  |  |  |  |  |  |  |  |
| **Past Medical History** | x |  |  |  |  |  |  |  |  |  |
| **Vital Signs** | x | x | x | x | x | x | x | x |  |  |
| <sup>§</sup> **Echocardiogram (LVEF)** | § |  |  | § | § | § | § |  |  |  |
| **SOFA-2** |  | x |  |  |  | x | x | x |  |  |
| **APACHE-II** |  | x |  |  |  | x | x | x |  |  |
| **ARDS Assessment** | x | x |  |  |  |  |  |  |  |  |
| **Complete Blood Count** | * |  |  | x |  | x | x | x |  |  |
| **Basic Metabolic Panel** | * |  |  |  |  | * | * | * |  |  |
| **Hepatic Panel** | * |  |  |  |  | * | * | * |  |  |
| **Troponin-T** | * |  |  |  |  | * | * |  |  |  |
| **hCG** | * |  |  |  |  |  |  |  |  |  |
| **Coagulation Panel** | * |  |  |  |  |  |  |  |  |  |
| **Arterial Blood Gases** | * | * |  | * | * | * | * | * |  |  |
| **Venous Blood Gases** | * | x |  | x | x | x | x | x |  |  |
| **Lactate** | * | x |  | x | x | x | x | x |  |  |
| † **Research Blood Drawn** |  | x |  | x | x | x | x | x |  |  |
| **Intervention** |  | x | x | x | x | x | x |  |  |  |
| **Outcome Assessment** |  |  |  |  |  | x | x | x | x | x |
| **Safety Outcome Assessment** |  |  | x | x | x | x | x | x | x | x |

<sup>‡</sup> Hour 0 marks the initiation of the intervention protocol; ‡ Vital signs will be recorded hourly throughout the trial; <sup>§</sup> LVEF will be obtained from TTE or TEE echocardiography, using the most recent study prior to hospitalization, as well as any echocardiograms performed during the current hospitalization; \*Labs denoted with astericks will be recorded if available; labs denoted with x are research blood draws; †Research Blood Drawn information is listed in table 3.
Abbreviations: ABG: Arterial Blood Gases; VBG: Venous Blood Gases; SOFA-2: Sequential Organ Failure Assessment; APACHE II: Acute Physiologic Assessment and Chronic Health Evaluation; ARDS Assessment: Acute Respiratory Distress Syndrome; hCG: Human chorionic gonadotropin; LVEF: Left ventricular ejection fraction; TTE: transthoracic echocardiography; TEE Transesophageal echocardiography.

### Sample size

Power and sample size calculations are based on the distribution of renin levels found in prior analysis of the VItamin C, Thiamine And Steroids in Sepsis (VICTAS) cohort.^19^ At a two-sided type 1 error rate of 5%, there will be 90% power to detect a 25% difference in renin at 24 hours with a target enrollment of n=78 septic shock patients (n=39 in each group); and 80% power to detect a 21.7% difference. To account for potential withdrawals after randomization, inclusion of participants who may later be found not to meet eligibility criteria, clinical improvement, or death, the study will enroll n=100 patients in total, with n=50 participants in each group.

### Recruitment

Recruitment is planned to start in November 2026. Data collection is expected to be completed by July 2030.

## METHODS: Assignment of Interventions

### Randomization

#### Sequence Generation, Allocation concealment and Implementation

Randomization will be completed in permuted blocks of varying sizes stratified by study-site. Allocation sequence will be computer-generated with a 1:1 ratio. Allocation concealment will be ensured using a secure, centralized electronic randomization system. The randomization sequence will be implemented through the REDCap randomization module, which will automatically assign participants to an intervention after eligibility confirmation and confirmation of minimum required NED. Investigators responsible for participant enrollment will not have access to the allocation sequence.

#### Blinding

This study is an open-label trial. Blinding is not implemented because it may be important for clinical decision-making regarding adjuvant or rescue vasopressor selection to know the study vasopressor that subjects are receiving.

## METHODS: Data Collection, Management and Analysis

### Data Collection Methods

Clinical, biological, and laboratory data will be collected at baseline and at predefined study timepoints, including 0, 3, 12, 24, 48 and 72 hours. Data will include demographics, comorbidities, clinical variables, disease severity scores (APACHE-II, SOFA-II), daily fluid balance, laboratory values, biomarker measurements, and outcome measures specified in the study protocol. Hourly measurements of vital signs and vasopressor use will be obtained throughout the intervention period. **Table 2 (Participant Timeline) and Table 3 (below)** show an overview of the planned data collection, sampling timepoints, and schedule for clinical and research blood analysis.

**Table 3.**
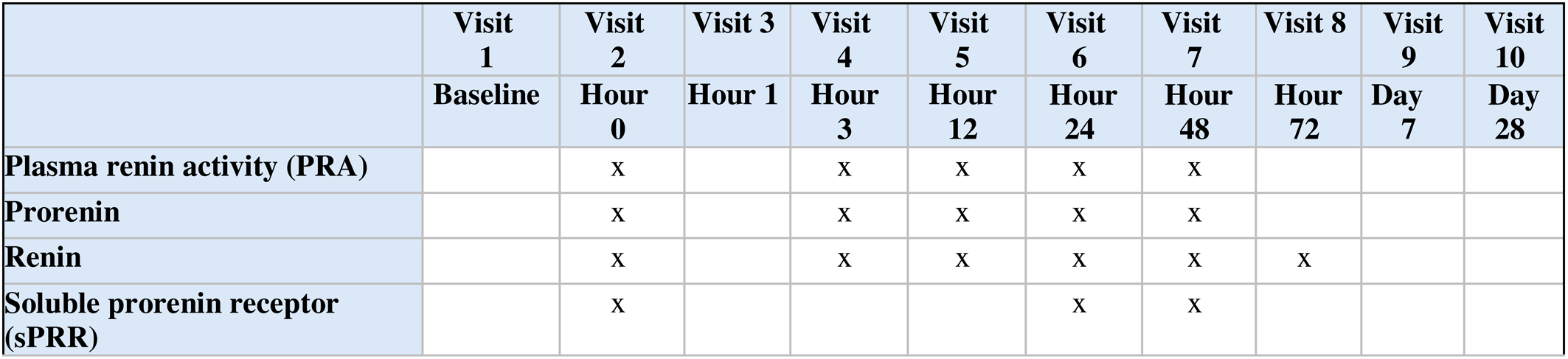
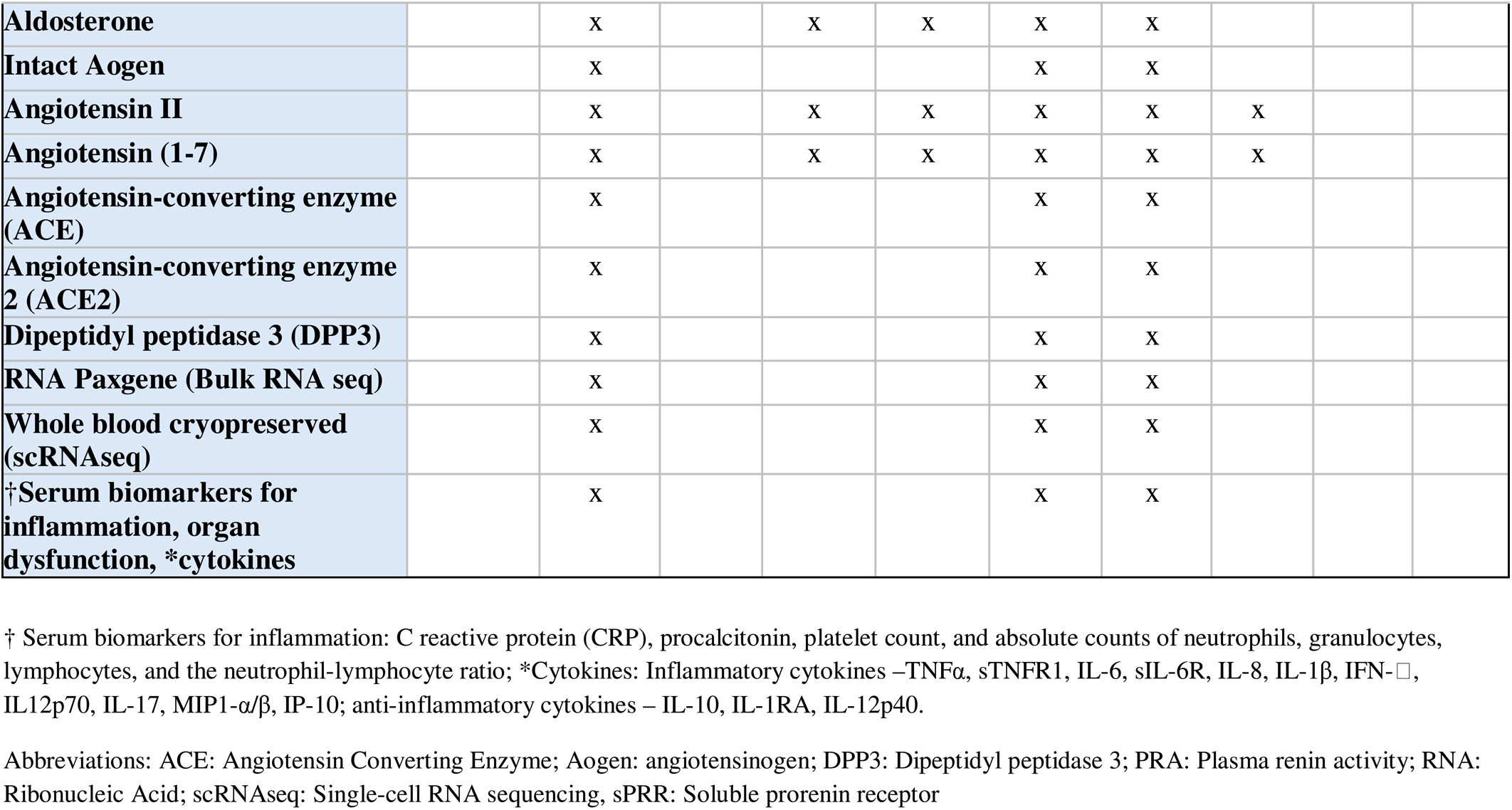
Research Blood Analysis.

|  | Visit 1 | Visit 2 | Visit 3 | Visit 4 | Visit 5 | Visit 6 | Visit 7 | Visit 8 | Visit 9 | Visit 10 |
|---|---|---|---|---|---|---|---|---|---|---|
|  | **Baseline** | **Hour 0** | **Hour 1** | **Hour 3** | **Hour 12** | **Hour 24** | **Hour 48** | **Hour 72** | **Day 7** | **Day 28** |
| **Plasma renin activity (PRA)** |  | x |  | x | x | x | x |  |  |  |
| **Prorenin** |  | x |  | x | x | x | x |  |  |  |
| **Renin** |  | x |  | x | x | x | x | x |  |  |
| **Soluble prorenin receptor (sPRR)** |  | x |  |  |  | x | x |  |  |  |
| **Aldosterone** |  | x |  | x | x | x | x |  |  |  |
| **Intact Aogen** |  | x |  |  |  | x | x |  |  |  |
| **Angiotensin II** |  | x |  | x | x | x | x | x |  |  |
| **Angiotensin (1-7)** |  | x |  | x | x | x | x | x |  |  |
| **Angiotensin-converting enzyme (ACE)** |  | x |  |  |  | x | x |  |  |  |
| **Angiotensin-converting enzyme 2 (ACE2)** |  | x |  |  |  | x | x |  |  |  |
| **Dipeptidyl peptidase 3 (DPP3)** |  | x |  |  |  | x | x |  |  |  |
| **RNA Paxgene (Bulk RNA seq)** |  | x |  |  |  | x | x |  |  |  |
| **Whole blood cryopreserved (scRNAseq)** |  | x |  |  |  | x | x |  |  |  |
| **†Serum biomarkers for inflammation, organ dysfunction, \*cytokines** |  | x |  |  |  | x | x |  |  |  |

† Serum biomarkers for inflammation: C reactive protein (CRP), procalcitonin, platelet count, and absolute counts of neutrophils, granulocytes, lymphocytes, and the neutrophil-lymphocyte ratio; \*Cytokines: Inflammatory cytokines –TNFα, sTNFR1, IL-6, sIL-6R, IL-8, IL-1β, IFN-□, IL12p70, IL-17, MIP1-α/β, IP-10; anti-inflammatory cytokines – IL-10, IL-1RA, IL-12p40.
Abbreviations: ACE: Angiotensin Converting Enzyme; Aogen: angiotensinogen; DPP3: Dipeptidyl peptidase 3; PRA: Plasma renin activity; RNA: Ribonucleic Acid; scRNAseq: Single-cell RNA sequencing, sPRR: Soluble prorenin receptor

Data will be obtained from the electronic medical record, and study-specific assessments. For measurement of the biological outcomes, up to 28 mL of blood per timepoint will be drawn and will be transported to laboratories at Atrium Health-Wake Forest Baptist and Massachusetts General Hospital for analysis of RAAS peptides and immune function studies, respectively.

### Data collection and management

Clinical data will be recorded by trained research study staff through REDCap, a secure, password-protected, HIPAA-compliant, IRB-approved, electronic data capture (EDC) system. Access to the database will be restricted to authorized study personnel. Each participant will be assigned a unique identifier number to maintain confidentiality, and identifiable information will be stored separately from study data. Biological samples will be stored in secured laboratory facilities following institutional biospecimen storage guidelines.

Data quality will be reviewed through automated validation rules within REDCap and routine data monitoring and verification procedures to ensure accuracy and completeness of the dataset.

### Statistical Methods

#### Statistical Analysis

Intention-to-treat principles will be used for all analyses unless otherwise stated. Categorical measures will be summarized as frequency (percentage). Continuous measures will be summarized as both mean (standard deviation) and median (minimum, 25th percentile, 75th percentile, maximum). All data will be analyzed and visualized by graphical summaries such as boxplots and histograms to ensure approximate normal distribution with adequate fit for the appropriate statistical model, and deviations from assumptions will utilize appropriate transformations or modifications of modeling approach to ensure goodness of fit. All analyses will be conducted using either R (Vienna, Austria) or SAS (Cary, North Carolina) statistical software.

#### Primary analysis

The primary outcome will be analyzed using a linear mixed effects model for repeated measures with random intercepts and restricted maximum likelihood estimation assuming equivalence between Ang-II and NE groups under the null hypothesis. The model for renin will include treatment (Ang-II vs. NE), time (0, 3, 12, and 24h), and treatment-by-time interaction fixed effects. The primary estimand of interest is represented by the treatment-by-time interaction effect, interpreted as the difference in renin response to treatment between groups from baseline to 24 hours, i.e., renin trajectory, with subsequent contrast in 24-hour renin levels to determine differences between groups.

Models will be additionally adjusted for disease severity (e.g., APACHE-II score) at randomization and study site. Although participants will be randomized, we will assess for between-group differences in baseline demographic and clinical characteristics, and the model will be further adjusted for any differences detected.

The linear mixed effects model is robust to missing data (i.e., missing measured renin or attrition by death occurring prior to 24h) under a missing at random (MAR) assumption. However, among this patient population, missingness may plausibly be due to worsening symptoms over time related to renin or unobserved latent variables that could result in a violation of the MAR assumption. Therefore, to assess the robustness of results to attrition, we will conduct a subsequent sensitivity analysis as has been recommended.^20^ Specifically, we will adapt pattern mixture modeling^21^ in which results will be averaged over participants who survive and provide renin data to 24h and those who do not.

#### Secondary analyses

For secondary outcomes, general linear mixed models similar to the primary analysis will be used to determine longitudinal differences in continuous biomarkers and clinical measures between Ang-II and NE groups. Severity of illness scores will be treated as ordinal variables. Vasopressor free hours (VFH) and organ failure free days (OFFD) will be treated as ordinal variables and interpreted as a count of hours or days. Both VFH and OFFD will be coded as -1 for participants who die before 72 hours or 28 days, respectively. Thus, these outcomes can take any integer value between (a) -1 and 72 hours, or (b) -1 and 28 days. Differences in VFH and OFFD between treatment groups will be analyzed using proportional odds logistic regression due to expected bimodal distributions of these outcomes. The corresponding odds ratios will indicate the difference in odds between treatment groups for more hours alive and vasopressor-free, or days alive and free of vasopressors, RRT, and mechanical ventilation, respectively. Differences in safety outcomes between Ang-II and NE groups will be evaluated using logistic regression adjusted for the same covariates as described in the analysis of the primary outcome. All analyses, including the primary outcome of plasma-renin trajectory, will use a modified intention-to-treat approach as follows: all patients who are randomized to treatment will be included for analysis within the treatment group they were assigned to, regardless of whether they were missing measurements or died before the primary endpoint time of 24-hours, provided that the study protocol was actually started with the treatment assigned. Multiplicity among higher dimensional RAAS biomarker analyses will be controlled with a false discovery rate target of 0.05.^22^

## METHODS: Monitoring

### Data Monitoring Committee

The principal role of the Data and Safety Monitoring Board (DSMB) is to ensure the safety of patients in the MANTRA trial. The DSMB will regularly monitor trial data to assess study performance and safety, and make recommendations with respect to adverse events, interim study results including safety outcomes, and safety concerns. The DSMB will comprise 4 clinician scientists and 1 senior epidemiologist and data scientist of significant repute and operate independent of the study sponsor. The DSMB will be updated and will convene as needed after every 6 months during study enrollment and report on its findings.

### Trial Monitoring

The principal investigator will be responsible for the overall monitoring of the data and safety of study participants. Daily patient screening and enrollment, protocol compliance, data collection, and outcome assessment will be performed by trained study staff with investigator oversight. Study staff and investigators will be in close daily contact with the clinical team during the intervention period. Data quality will be reviewed remotely using front end range and logic checks at the time of data entry and back-end monitoring of data using analytic reports.

### Safety Monitoring

Each participating investigator has primary responsibility for the safety of the individual participants under his or her care. The safety monitoring plan will specify all items related to safety including adverse event (AE) reporting, AE grading and determination of relationship between the AE and the intervention. AE definitions and grading are outlined in **Appendix A**.

## ETHICS AND DISSEMINATION

### Research Ethics Approval

This clinical trial involving humans will be conducted in accordance with Good Clinical Practice (GCP) guidelines and the principles of the Declaration of Helsinki. The study protocol was approved by the Advocate Health-Wake Forest School of Medicine ethics committee and is registered on clinicaltrials.gov (NCT06746753).

### Plans for communicating important protocol modifications

Any amendments to the study protocol will be documented and submitted to Institutional Review Boards IRB00115287 and communicated in a timely manner to all relevant parties to ensure consistent implementation across all study sites.

### Consent or assent

Consent will be obtained by a licensed physician investigator trained on the study. The consenting investigator is responsible for ensuring that the patient and/or the subject’s LAR understand the risks and benefits of participating in the study. The IRB-approved informed consent document will be used to explain the objective of the study, risks and benefits in simple terms before the patient is entered into the study. Subjects for whom consent was initially obtained from a LAR, but who subsequently regain decision-making capacity while in the hospital, will be approached for consent to participation, including use of collected data and continuation of study procedures.

### Confidentiality

All participant personal information, as well as all laboratory specimens, will be collected and stored securely with unique study identifiers. Access will be limited to authorized study personnel, and data will be encrypted for electronic storage. Identifiable information will be removed or anonymized for analysis, publication, or archiving with the purpose of maintaining confidentiality before, during, and after the trial.

## Supporting information

Response to Reviewers

## Data Availability

Data will be available from the corresponding author on reasonable request.

## AUTHOR’S CONTRIBUTION

Michael R. Filbin, Daniel E. Leisman, Marcia B.Goldberg, Ashish K.Khanna, Mark C. Chappell and Clark Files conceived and designed the study. Michael R. Filbin, Daniel E. Leisman, Ana Pachano-Bravo, Marcia B. Goldberg, Kathryn Hibbert, Simon A. Mahler, Ryan C. Maves, Andrew Petrilli, D. Clark Files, Kevin Gibbs, Mark C. Chappell, Ashish K. Khanna, and other study investigators contributed to study development, interpretation of data, and critical revision of the manuscript.

Ana Pachano-Bravo, Olivia Nelson, Benjamin Rappaport, Lynnette Harris, Estelle B. Besong, and Brandon Reeves contributed to participant enrollment, data acquisition, data management, and study operations.

Christopher L. Schaich and Mark C. Chappell contributed to laboratory methods, assay development, and/or data analysis. Christopher L. Schaich performed the primary statistical analyses. Michael R. Filbin, Ashish K.Khanna and Daniel E. Leisman drafted the initial manuscript.

All authors contributed substantially to data interpretation, critically reviewed and revised the manuscript for important intellectual content, approved the final version of the manuscript, and agree to be accountable for all aspects of the work.

## PATIENT AND PUBLIC INVOLVEMENT STATEMENT

Patients were not involved in the development of this research protocol.

