## Appendix A for "Mechanistic Assessment of Norepinephrine Therapy versus Angiotensin-II on Organ Function in Septic Shock (MANTRA): Study Protocol for a multicenter open-label randomized trial"

**Definitions and Grading of Adverse Events**

Definition of Adverse Events and Serious Adverse Event

Adverse events (AEs) are defined as any unfavorable and unintended sign, symptom, or disease temporally associated with the study. Abnormal laboratory findings are not considered AEs in this study. Only CTCAE grade 3 or higher AEs will be recorded in this study from randomization through study day 7 or Hospital discharge, whichever occurs first.

Serious Adverse Events (SAEs) are defined as any AE that results in fatal, life-threatening, permanently or substantially disabling, requires or prolongs hospitalization. All serious AEs will be recorded from randomization through study day 7, whichever occurs first. Based on the guidelines of the International Conference on Harmonization (ICH), SAEs must be reported within 24 hours to Wake Forest CC, regardless of the investigator's opinion of causation.

Protocol-defined anticipated clinical events (PDACEs) of sepsis are clinical events expected to occur commonly in the target population even in the absence of study interventions. PDACEs (CTCAE grade 3 or higher) will be considered clinical outcomes rather than AEs, unless deemed related to the administration of study drug or conduct of study procedures. CTCAE grade 3 or higher PDACEs deemed related to the study drug will be recorded as AEs. PDACEs are as follows:

- Cardiovascular events: New or escalating medical management (addition of new vasopressor, inotrope, anti-arrythmic therapy) or electrical cardioversion for treatment of worsening hypotension or cardiac arrhythmia. New myocardial infarction.
- Respiratory events: Worsening respiratory condition defined as receipt of new high flow nasal oxygen (HFNO), non-invasive mechanical ventilation, invasive mechanical ventilation, or extracorporeal membrane oxygenation (ECMO).
- Hepatic events: Worsening hepatic function defined as new or worsening ascites, gastrointestinal bleeding, hepatic encephalopathy.
- Renal events: New requirement for renal replacement therapy.
- Neurological events: New delirium, transient ischemic attack or cerebrovascular event.
- Hematologic events: New arterial or venous thromboembolic event excluding superficial thrombophlebitis, major bleeding requiring >2 units of blood within 24 hours, disseminated intravascular coagulation (DIC).
- Infection events: New infection requiring antimicrobial treatment.
- Death

Of note, the protocol-specified safety outcomes including venous thromboembolism and onset of arrythmia will be classified as PDACEs. If a patient's treatment is discontinued as a result of an AE or PDACE, site personnel will report this in an adverse event case report form.

Severity of an Adverse Event

Throughout the study, the investigator will determine whether any CTCAE grade 3 or higher AEs have occurred. To define severity, the Common Terminology Criteria for Adverse Events (CTCAE) v6.0 will be used.^23^ The CTCAE grades severity from 1 to 5. (Grade 1: Mild AE, Grade 2: Moderate AE, Grade 3: Severe AE Grade 4: Life-Threatening or Disabling AE, Grade 5: Death related to AE.

Causality of an Adverse Event

To determine relatedness, the site PI or Co-PI will determine how likely it is that the AE is caused by the study intervention. The study uses the following AE attribution scale:

- **Definitely:** The AE is clearly related to the intervention. It must meet all 3 of these conditions:
  - Has a reasonable temporal relationship to intervention.
  - Could not possibly have been produced by the participant’s clinical state or other interventions.
  - Follows a known pattern of response to intervention
- **Probable:** The AE is likely related to the intervention and might meet 2 of the 3 conditions detailed above.
- **Possible:** The AE may be related to the intervention and might meet 2 of the 3 conditions detailed above.
- **Unlikely:** The AE is doubtfully related to the intervention and might meet 2 of the following conditions:
  - Does not follow a known pattern of response to intervention
  - Could have been produced by the participant’s clinical state or other interventions.
- **Unrelated:** The AE is clearly not related to the intervention; its temporal relationship between treatment exposure and AE is incompatible, and/or AE is clearly due to different causes such as underlying disease.
